# Orthostatic Blood Pressure Transitions: Association with Symptoms and Frailty

**DOI:** 10.1101/2025.09.17.25336032

**Authors:** Edmur C. A. Filho, Fernando Yue Cesena, Bruno N. Blaas, Magno M. P. de Faria, Sarah T. de Biaso, Caroline Y. Yoshioka, Mário S. C. Pires, Rogerio M. C. Britto, Marcio G. de Sousa, Fernanda M. Consolim-Colombo, Jonathan B. Souza, Antônio Gabriele Laurinavicius

## Abstract

**Objective:** Individuals with hypertension may transition between different orthostatic blood pressure (BP) phenotypes within minutes. We studied the temporal patterns of orthostatic BP responses and their relationship to symptom development in older hypertensive individuals with and without frailty.

**Design and method:** A cross-sectional study was conducted including patients aged ≥60 years. BP was measured seated and at 1- and 3-minutes post-standing. Orthostatic hypotension (OH) and orthostatic hypertension (OHT) were defined in accordance with current guidelines. Orthostatic intolerance (OI) was defined as symptoms upon standing. Frailty was assessed using the Clinical Frailty Scale. Multivariable logistic regression models evaluated associations between orthostatic BP phenotypes, OI and Frailty.

**Results:** We included 461 hypertensive adults (mean age: 72.5 ± 7.0 years, 70% female). The prevalence of OH and OHT was 11% and 10%, respectively. About 50% of individuals with OH or OHT at minute 1 normalized their BP by minute 3, while a similar absolute number with normal BP at minute 1 developed OH or OHT by minute 3. Frail individuals exhibited a twofold higher prevalence of OH (OR 2.39, p = 0.023), and a more than threefold higher prevalence of OHT (OR 3.60, p < 0.001). In fully adjusted models, OI was associated with both systolic OH (OR 3.05, p = 0.019) and OHT (OR 2.33, p = 0.041).

**Conclusions:** Orthostatic BP phenotypes in hypertensive older adults were dynamic, with frequent shifts between 1 and 3 minutes of standing. Frailty and OI were strongly associated not only with OH but also with OHT.

## Introduction

Under physiological conditions, postural changes induce only minimal and transient effects on blood pressure (BP), due to prompt compensatory autonomic and hemodynamic responses.^1^ However, when cardiovascular reactivity is impaired - often in the context of aging, multimorbidity, or autonomic dysfunction - these compensatory mechanisms may be blunted or delayed, leading to abnormal BP fluctuations upon standing.

In clinical practice, two abnormal orthostatic BP phenotypes are widely recognized: orthostatic hypotension (OH) and orthostatic hypertension (OHT). Both are considered markers of autonomic dysregulation and have been independently associated with adverse cardiovascular outcomes, including stroke and myocardial infarction, as well as falls and all-cause mortality.^1,2,3^ These phenotypes frequently coexist with hypertension and may be exacerbated by antihypertensive pharmacotherapy, posing significant challenges to optimal BP control, particularly in frail older adults.^4,5^ Orthostatic intolerance (OI) - defined as the presence of symptoms such as dizziness, lightheadedness, blurred vision, or near-syncope within three minutes of standing - is most frequently associated with OH, but may also occur in the context of OHT.^8^

Both OH and OHT appear to be more prevalent and pronounced in frail individuals,^9,10,11^ likely reflecting mechanisms such as autonomic dysfunction, increased arterial stiffness, polypharmacy, and impaired baroreflex sensitivity.^4,12^ Consequently, OI is also more frequently observed in this population and may further contribute to adverse outcomes, including falls, functional decline, and hospitalization.^13^

Given the dynamic nature of BP regulation, individuals may transition between orthostatic BP phenotypes within minutes, and measurements at different time points may yield discrepant results, interfering with phenotype classification. Accordingly, current guidelines recommend measuring BP at both 1 and 3 minutes after standing.^14,15^ The temporal patterns of orthostatic BP responses and their relationship to symptom development, however, remain poorly understood. This variability may have important clinical implications for risk stratification and for elucidating the mechanisms underlying orthostatic intolerance. The role of frailty in modulating these short-term BP dynamics is also unclear.

This study aimed to: (1) characterize orthostatic BP changes from minute 1 to minute 3 after standing in older individuals with hypertension; (2) investigate the association between clinical frailty and orthostatic BP phenotypes; and (3) examine the relationship between OI and orthostatic BP phenotypes in this population.

## Methods

### Study design and population

This cross-sectional study included patients attending a tertiary outpatient hypertension clinic at the Instituto Dante Pazzanese de Cardiologia, São Paulo-SP, Brazil, between April and June 2024. Individuals aged ≥60 years were recruited through convenience sampling and underwent assessment of BP, frailty status, and OI. Patients with fewer than three seated BP measurements or missing BP data at 1 or 3 minutes after standing were excluded. The study was conducted in accordance with institutional and international ethical standards, and written informed consent was obtained from all participants prior to enrollment.

### Frailty assessment

The Clinical Frailty Scale (CFS) was employed following the current recommendations and patients were stratified into two groups based on their CFS scores.^15^ Those with a score of less than 5 were classified as non-frail, while those with a score of 5 or more were considered frail.

### Blood pressure measurements

BP was measured using an oscillometric device (Omron HBP-1120) in both seated and standing positions, following a standardized protocol. After at least five minutes of rest, three seated BP measurements were obtained; the average of the last two readings was used for analysis. Orthostatic BP was assessed at 1 and 3 minutes after standing from the seated position to evaluate postural BP changes. All measurements were performed by the attending physician.

### Orthostatic intolerance

OI was defined as at least one symptom such as palpitation, weakness, blurred vision, or dizziness upon standing.

### Orthostatic blood pressure phenotypes

OH was defined as a decrease in systolic blood pressure (SBP) of at least 20 mmHg or in diastolic blood pressure (DBP) of at least 10 mmHg within 1 or 3 minutes of standing.^6^ Systolic OH was defined as a reduction of ≥ 20 mmHg in SBP, and diastolic OH as a reduction of ≥ 10 mmHg in DBP. OHT was defined as an increase in SBP of at least 20 mmHg within the same timeframe, accompanied by a standing SBP of at least 140 mmHg.^7^

### Other variables

Data on medication use, history of stroke, diabetes mellitus (DM) or ischemic heart disease (IHD), height, weight, and smoking status were collected through patient interviews conducted by the attending physician. Laboratory and clinical data were extracted from patients’ medical records following the analysis of laboratory and imaging tests.

## Statistical analyses

Categorical variables are reported as the absolute and relative number of observations. Continuous variables are shown as mean ± standard deviation if normally distributed or median (interquartile range) if non-normally distributed. Normality was assessed by visual inspection of histograms and Q-Q plots.

The chi-squared test and, when appropriate, the Fisher’s exact test were used to compare categorical variables between non–frail and frail individuals. Continuous variables were compared by the Student’s t-test and, when equal variances between groups could not be assumed (Levene’s test), the Welch’s t-test.

Sankey diagrams were constructed to visualize changes in orthostatic BP phenotypes from 1 minute to 3 minutes after standing. Associations between frailty or OI and BP orthostatic phenotypes were evaluated by logistic regression. In these models, the dependent variable was the orthostatic BP phenotype (orthostatic systolic hypotension, orthostatic diastolic hypotension, or OHT), and the independent variable was the categorized CFS or OI. Three models were performed: Model 1, unadjusted; Model 2, adjusted for sex and age; and Model 3, further adjusting for sitting blood pressure (SBP when the outcome was orthostatic systolic hypotension or OHT, DBP for the association with orthostatic diastolic hypotension), and number of antihypertensive medications (categorized in ≥3 or <3) on top of Model 2.

The R software (version 4.4.2) and the jamovi software (version 2.6.13.0)^3^ were used for data manipulation and statistical analyses. A p-value <0.05 was considered statistically significant for all analyses.

## Results

### Study sample and participant characteristics

Figure 1 shows the flowchart of included and excluded participants. Compared to non-frail individuals, frail participants were older, had a higher prevalence of diabetes, a higher proportion of previous stroke, and more frequently reported OI (Table 1).

**Table 1.** Participant characteristics according to frailty status

| Characteristic | Total<br>(n=461) | Non-frail (CFS <5,<br>n=413) | Frail (CFS ≥5,<br>n=48) | p-value |
| --- | --- | --- | --- | --- |
| Female sex | 322 (70%) | 286 (69%) | 36 (75%) | 0.411 |
| Age (years) | 72.5 ± 7.0 | 72.0 ± 6.7 | 77.3 ± 8.0 | <0.001 |
| Diabetes mellitus | 266 (58%) | 232 (56%) | 34 (72%) | 0.035 |
| Previous stroke | 28 (6%) | 21 (5%) | 7 (15%) | 0.008 |
| Previous coronary artery disease* | 46 (12%) | 42 (12%) | 4 (11%) | 1.000 |
| Smoking |  |  |  |  |
| - Never | 295 (65%) | 256 (63%) | 39 (83%) |  |
| - Previous | 136 (30%) | 130 (32%) | 6 (13%) | 0.013 |
| - Current | 25 (5%) | 23 (5.6%) | 2 (4.3%) |  |
| Number of antihypertensive medications | 3.5 ± 1.3 | 3.5 ± 1.3 | 3.3 ± 1.3 | 0.282 |
| Orthostatic intolerance | 49 (11%) | 40 (10%) | 9 (20%) | 0.048 |
| Body mass index (kg/m<sup>2</sup>) | 28.9 ± 5.4 | 29.0 ± 5.3 | 28.2 ± 6.2 | 0.316 |

Data shown as n (%) or mean±standard deviation.
\* Data available for 385 (84%) participants.
CFS: Clinical Frailty Scale.

**Figure 1.**
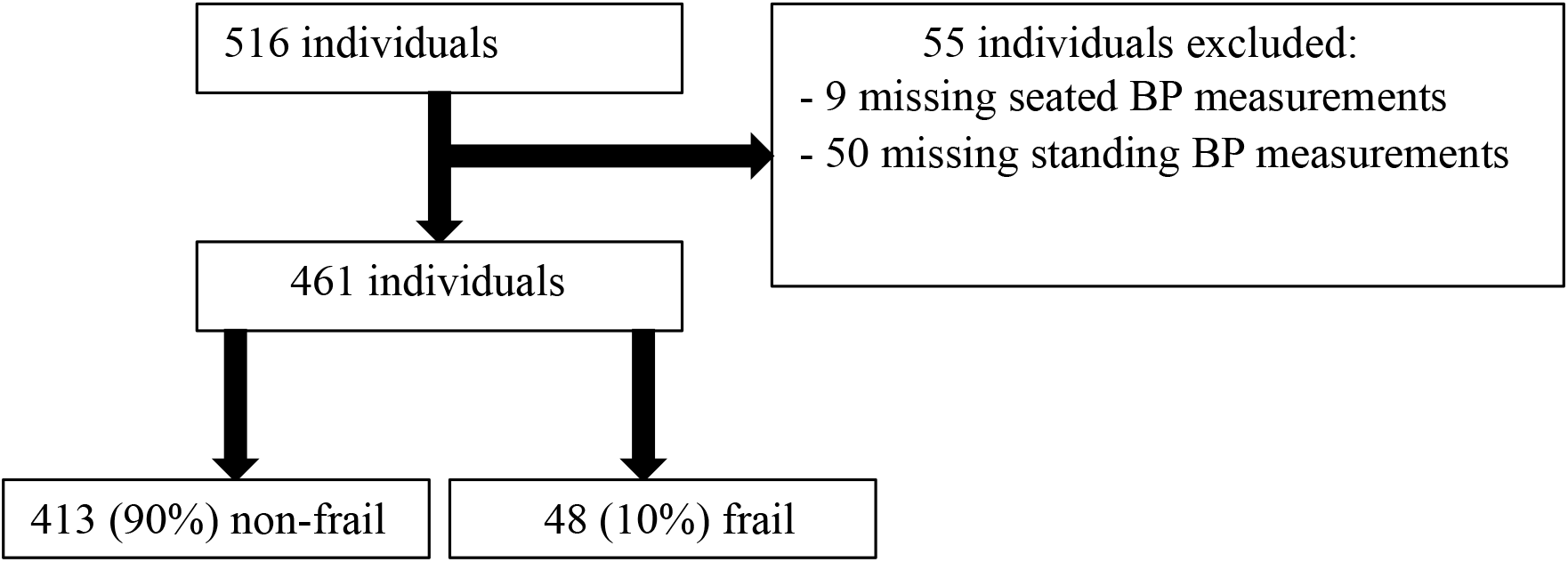
Flowchart depicting included and excluded individuals. CFS: Clinical Frailty Scale.

### Orthostatic blood pressure changes from 1 to 3 minutes of orthostasis

Overall, the prevalence of OH and OHT was 11% and 10%, respectively. Compared to non-frail participants, frail individuals exhibited a twofold higher prevalence of OH (21% vs. 10%, OR 2.39, 95% confidence interval [CI] 1.11 to 5.14, *p* = 0.023, Table 2) and a threefold higher prevalence of OHT (25% vs. 9%, OR 3.60, 95% CI 1.72 to 7.54, *p* < 0.001, Table 2).

**Table 2.** Blood pressure measurements at sitting and standing positions and orthostatic phenotypes according to frailty status.

| Parameter | Total<br>(n=461) | Non-frail (CFS<br><5, n=413) | Frail (CFS<br>≥5, n=48) | p-value |
| --- | --- | --- | --- | --- |
| Sitting SBP (mmHg) | 150 ± 23 | 149 ± 22 | 156 ± 24 | 0.028 |
| Sitting DBP (mmHg) | 79 ± 12 | 78 ± 12 | 80 ± 14 | 0.264 |
| 1-minute standing SBP (mmHg) | 151 ± 24 | 150 ± 24 | 160 ± 25 | 0.008 |
| 1-minute standing DBP (mmHg) | 84 ± 14 | 83 ± 13 | 85 ± 16 | 0.307 |
| Change in SBP from sitting to 1-minute standing (mmHg)* | -1.6 ± 12.7 | -1.4 ± 12.1 | -3.5 ± 17.2 | 0.417** |
| Change in DBP from sitting to 1-minute standing (mmHg)* | -4.9 ± 8.7 | -4.9 ± 8.4 | -5.0 ± 11.0 | 0.942** |
| 3-minute standing SBP (mmHg) | 152 ± 24 | 151 ± 23 | 161 ± 24 | 0.005 |
| 3-minute standing DBP (mmHg) | 85 ± 13 | 84 ± 13 | 87 ± 16 | 0.107 |
| Change in SBP from sitting to 3-minute standing (mmHg)* | -2.3 ± 12.6 | -2.1 ± 12.2 | -4.5 ± 15.6 | 0.318** |
| Change in DBP from sitting to 3-minute standing (mmHg)* | -5.9 ± 8.1 | -5.8 ± 7.7 | -6.9 ± 10.7 | 0.468** |
| Systolic orthostatic hypotension | 31 (7%) | 27 (7%) | 4 (8%) | 0.550 |
| Diastolic orthostatic hypotension | 27 (6%) | 20 (5%) | 7 (15%) | 0.007 |
| Systolic or diastolic orthostatic hypotension | 51 (11%) | 41 (10%) | 10 (21%) | 0.023 |
| Orthostatic hypertension | 47 (10%) | 35 (9%) | 12 (25%) | <0.001 |

Data shown as n (%), mean±standard deviation
\* Change in BP means sitting BP minus standing BP
\*\* Welch's t-test
BP: blood pressure; CFS: clinical frailty scale; DBP: diastolic blood pressure; SBP: systolic blood pressure.

Figure 1 presents transitions in orthostatic BP phenotypes between 1 and 3 minutes of standing, stratified by frailty status. Among the 413 non-frail participants, the prevalence of OH and OHT at 1-minute post-standing was 7% (n = 28) and 5% (n = 19), respectively. One participant (0.2%) exhibited concurrent systolic OHT and diastolic OH. Among individuals with a normal BP response at 1 minute (88%), a number transitioned to OH (3%) or OHT (4%) by minute 3. Notably, 46% of those with OH at 1 minute showed normalization of BP at 3 minutes. Likewise, 42% of those with OHT at 1 minute exhibited a normal BP response at 3 minutes (Figure 1A).

Among the 48 frail participants, 19% (n = 9) presented OH and 14.6% (n = 7) OHT at 1 minute after standing (Figure 1B). Among those with a normal BP response at 1 minute, 3% became hypotensive and 12.5% hypertensive by 3 minutes. Of those with OH at 1 minute, 56% remained hypotensive, 33% normalized their BP response, and 11% (n = 1) transitioned to OHT at 3 minutes. Among participants with OHT at 1 minute, 43% remained hypertensive, while 57% returned to a normal BP response at 3 minutes (Figure 1B).

**Figure 1.**
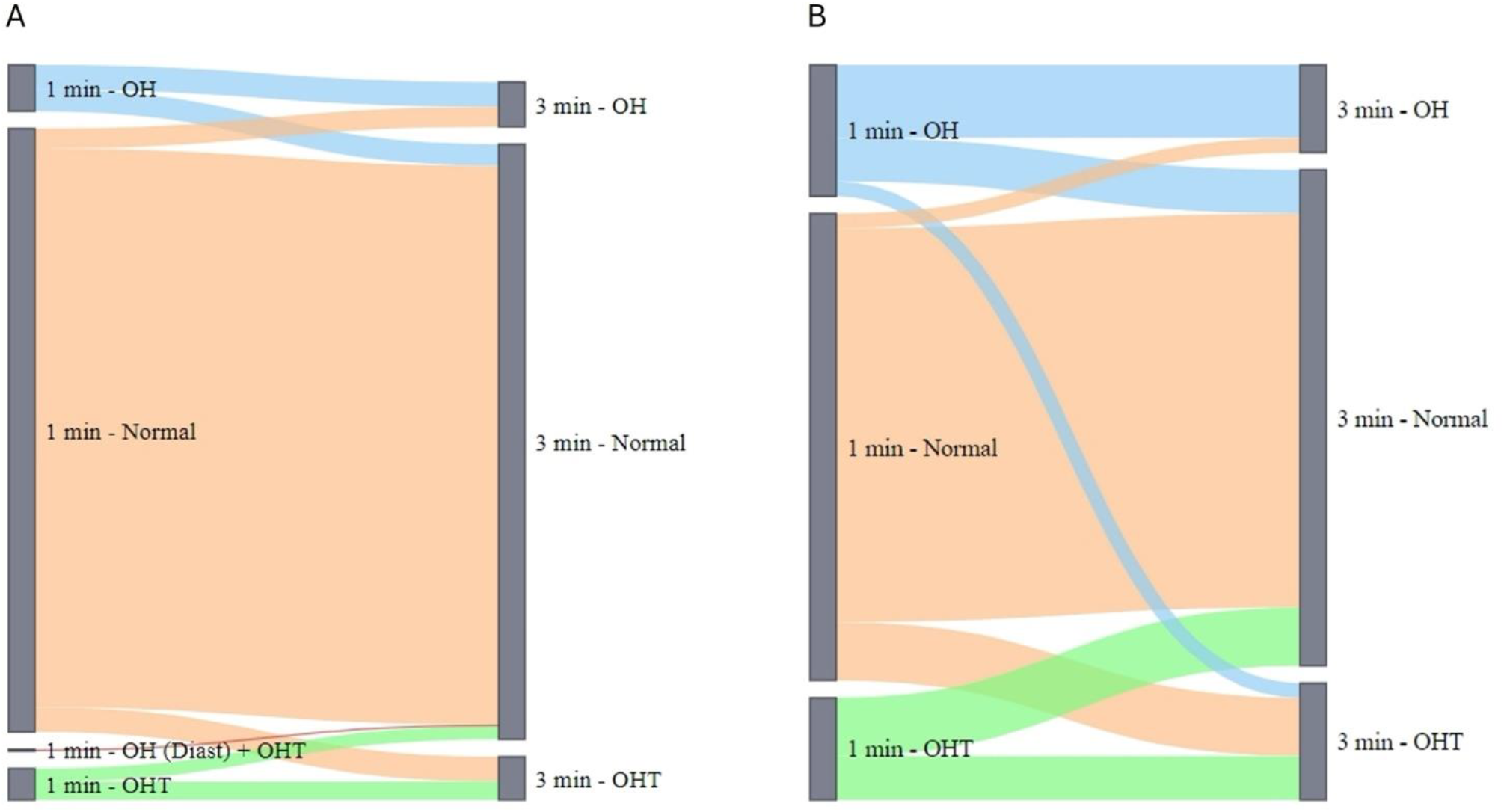
Changes in blood pressure orthostatic phenotypes between 1 minute and 3 minutes after standing in non-frail (CFS <5, n=413, **Panel A**) and frail individuals (CFS ≥5, n=48, **Panel B**). CFS: Clinical Frailty Scale; OH: orthostatic hypotension; OHT: orthostatic hypertension

### Association between frailty and orthostatic blood pressure phenotypes

Compared to non-frail participants, frail individuals exhibited higher systolic BP in both the sitting and standing positions, a higher prevalence of OH —driven primarily by a greater frequency of orthostatic diastolic drops— and an increased proportion of OHT. However, the mean absolute BP changes in response to standing did not differ significantly between the groups (Table 2).

In sex- and age-adjusted models, frailty was associated with diastolic OH, but not with systolic OH. The association between frailty and diastolic OH weakened after further adjustment for sitting diastolic BP and the number of antihypertensive medications (Figure 2). Frailty was also associated with OHT. In fully adjusted models, participants with a CFS ≥ 5 had a statistically significant 3.5-fold increase in the chance of presenting OHT (Figure 2).

**Figure 2.**
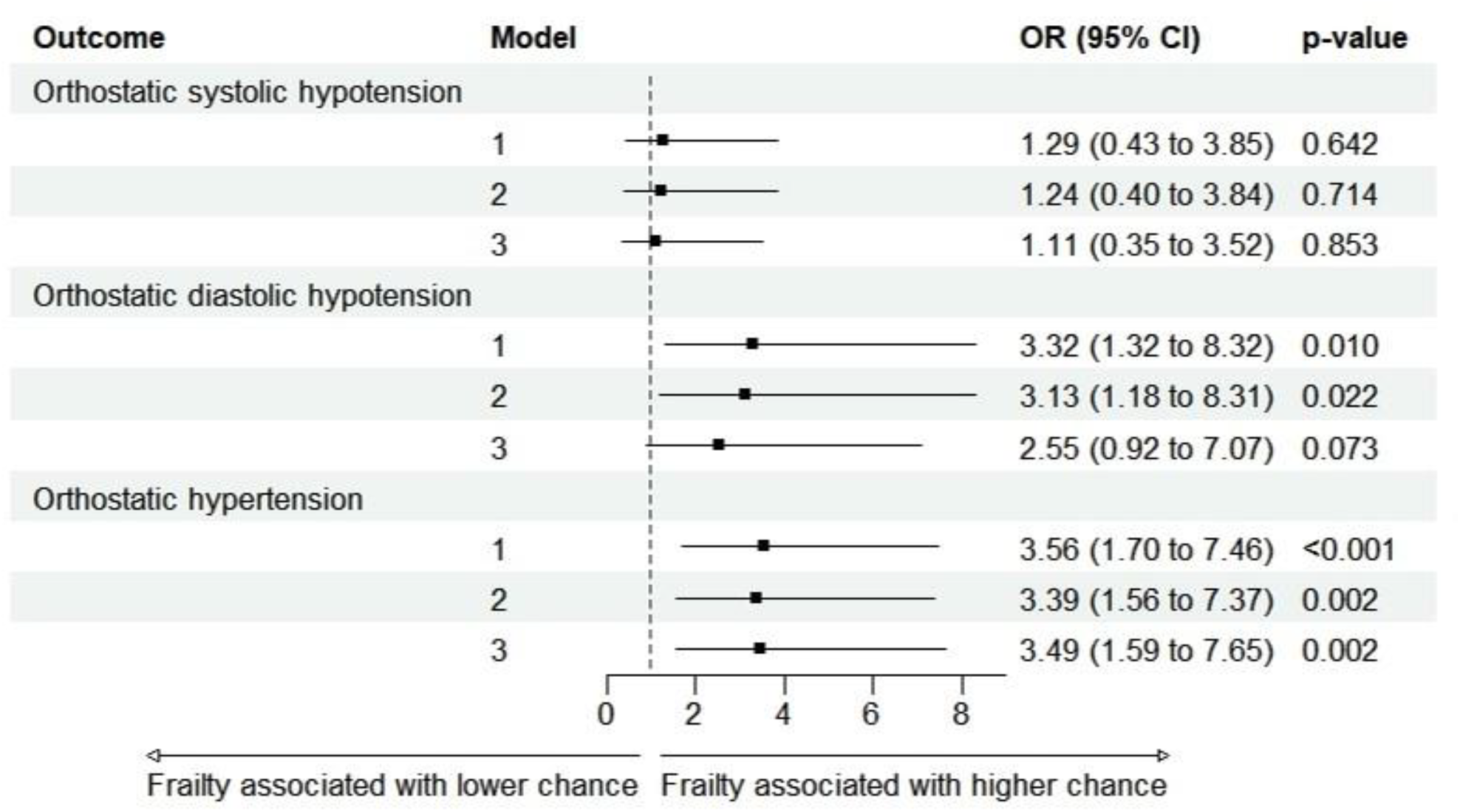
Association between frailty and blood pressure orthostatic phenotypes. The figure shows the ORs of a frailty status (CFS ≥5, in relation to CFS <5) being associated with each phenotype. Model 1: unadjusted; Model 2: adjusted for sex and age; Model 3: Model 2 further adjusted for sitting blood pressure and the number of antihypertensive medications. CFS: clinical frailty scale; CI: confidence interval; OR: odds ratio.

### Association between orthostatic intolerance and blood pressure phenotypes

Symptom data were available for 448 (97.2%) of the 461 participants included in the study. Symptoms were twice as common among frail compared to non-frail individuals (19.6% [9 of 46] vs. 10.0% [40 of 402], OR 2.20, 95% CI 0.99 to 4.89, *p* = 0.048). Moreover, OI was associated with a more than twofold higher chance of orthostatic systolic hypotension and OHT in unadjusted and adjusted models (Figure 3).

**Figure 3.**
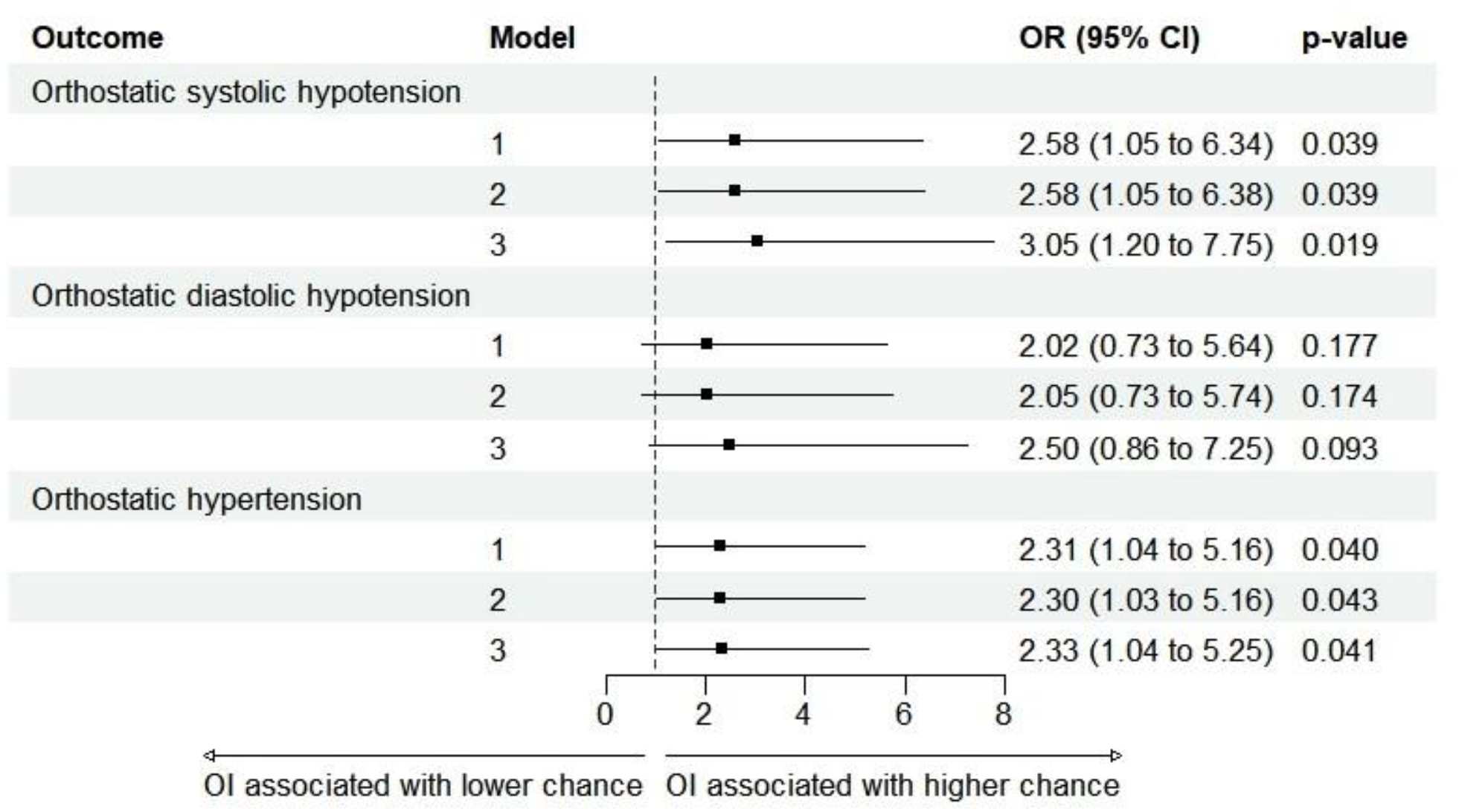
Association between OI and blood pressure orthostatic phenotypes. The figure shows the ORs of OI being associated with each phenotype. Model 1: unadjusted; Model 2: adjusted for sex and age; Model 3: Model 2 further adjusted for sitting blood pressure and the number of antihypertensive medications. CI: confidence interval; OI: orthostatic intolerance; OR: odds ratio.

## Discussion

Our study highlights the dynamic nature of BP responses during orthostasis, revealing substantial transitions in orthostatic BP phenotypes between 1 and 3 minutes after standing. In a hypertensive sample of older adults (mean age 72 years) attending a specialized hypertension center and receiving an average of 3.5 antihypertensive medications, the prevalence of OHT was comparable to that of OH, with each affecting approximately 10% of participants.

Notably, phenotype transitions between 1 and 3 minutes of standing were frequent, and relying on a single time point for BP measurement would have led to misclassification in nearly 50% of cases. These findings are consistent with recent data from Choi et al., who reported prevalences of 4.6% for OHT and 4.1% for OH in a similar older adult population in South Korea, based solely on BP measurements taken at 3 minutes post-standing.^12^

Approximately 10% of our study population met the Clinical Frailty Scale (CFS) criteria for frailty. As expected, frailty was associated with a higher likelihood of both OH and OHT, with a twofold increase in the prevalence of OH and a threefold increase in OHT compared to non-frail individuals. Although the mean absolute BP changes in response to standing did not differ significantly between frail and non-frail participants, phenotype shifts at 3 minutes were more pronounced in the frail group, driven primarily by a higher prevalence of delayed-onset OHT. These findings underscore that reliance on a single time point may fail to capture clinically relevant orthostatic BP changes, particularly among frail older adults.

Frailty has traditionally been linked to OH, whereas OHT has received comparatively less attention and remains underexplored in this context. In our analysis, however, frailty was more strongly associated with OHT, with a 3.5-fold increased likelihood in fully adjusted models. While the clinical implications of OHT in frail older adults are still being elucidated, our findings highlight the importance of capturing the full spectrum of orthostatic BP responses through sequential measurements rather than relying on a single time point.

In our sample, frailty was also significantly associated with OI, with nearly 20% of frail individuals reporting symptoms related to abnormal orthostatic BP responses - almost twice the prevalence observed among non-frail participants. Both systolic OH and OHT were significantly associated with OI, whereas diastolic OH was not. Although seemingly counterintuitive, the association between OI and OHT has been previously documented. In a study by Lee et al., 1,033 patients with OI underwent autonomic function testing, including head-up tilt, and 3.7% were found to have OHT.^8^ These individuals exhibited increased peripheral vascular resistance and sympathetic activation during tilt testing. The mechanisms underlying OI in individuals with OHT remain unclear; however, our findings suggest that dynamic transitions between orthostatic BP phenotypes may partially account for the presence of symptoms in this group.

Interestingly, among frail participants, OI was not significantly associated with any abnormal orthostatic BP phenotype. This suggests that, in this population, orthostatic symptoms may stem from other underlying conditions beyond BP dysregulation.

Several limitations should be acknowledged when interpreting our results. The cross-sectional design precludes causal inference, and the use of convenience sampling in a single tertiary hypertension centre may limit the generalizability of our findings to broader elderly populations. The relatively small number of frail individuals may also have reduced the statistical power to detect associations in subgroup analyses. Frailty was assessed using the CFS, and the cut-off used to define frailty was arbitrary; alternative thresholds or more comprehensive frailty assessments could yield different results. Although orthostatic intolerance and abnormal orthostatic BP responses were more frequent among frail individuals, this group also had a higher prevalence of diabetes (72% vs. 56%) and prior stroke (15% vs. 5%) compared to non-frail participants. These conditions may have contributed to autonomic and neurosensory dysfunction—or other forms of dysautonomia unrelated to orthostatic changes—that were nevertheless classified as orthostatic intolerance in this study. Furthermore, orthostatic symptoms were self-reported, introducing the potential for recall bias, especially among older or cognitively impaired participants. Finally, while we adjusted the analysis for the number of antihypertensive medications, we did not account for pharmacologic classes, which may exert differential effects on orthostatic BP responses.

In conclusion, orthostatic BP phenotypes in older adults with hypertension are highly dynamic, with frequent shifts between 1 and 3 minutes of standing. Frailty was strongly associated not only with OH but also with OHT, underscoring the need to move beyond single time-point measurements. Our findings highlight the clinical relevance of sequential orthostatic BP assessment, particularly in frail populations, to better characterize risk and guide management.

## Data Availability

The data that support the findings of this study are available from the corresponding author upon request.

## Notes

### Competing Interest Statement

The authors have declared no competing interest.

### Clinical Trial

This study does not report results of a clinical trial and was therefore not registered in ClinicalTrials.gov or any other trial registry.

### Funding Statement

This work was supported by institutional resources from the Instituto Dante Pazzanese de Cardiologia. No external funding was received.

### Author Declarations

This study was approved by the Research Ethics Committee of Instituto Dante Pazzanese de Cardiologia, São Paulo, Brazil, with a waiver of informed consent.

